# Predicting poor mental health amongst older Syrian refugees in Lebanon during the COVID-19 pandemic: a multi-wave longitudinal study

**DOI:** 10.1101/2023.12.22.23300447

**Authors:** Berthe Abi Zeid, Leen Farouki, Tanya El Khoury, Abla Sibai, Carlos F. Mendes de Leon, Marwan F. Alawieh, Zeinab Ramadan, Sawsan Abdulrahim, Hala Ghattas, Stephen J. McCall

## Abstract

**Background:** The COVID-19 pandemic has worsened pre-existing vulnerabilities among older Syrian refugees in Lebanon, potentially impacting their mental health. This study aimed to identify predictors of poor mental health amongst older Syrian refugees living in Lebanon during the pandemic.

**Methods:** This study used repeated cross-sectional data from a multi-wave telephone survey (September 2020-March 2022). It was conducted among Syrian refugees aged 50 years or older from households that received assistance from a humanitarian organization. Poor mental health was defined as a Mental Health Inventory-5 score of 60 or less. Its trend over time was assessed using growth curve model; and, its predictors were identified using wave one data, through backwards stepwise logistic regression. The model’s internal validation was conducted using bootstrapping.

**Findings:** There were 3,229 participants (median age=56 [IQR=53-62]) and 47.5% were female. At wave one, 76.7% had poor mental health, and this increased to 89.2% and to 92.7% at waves three and five, respectively (β=0·52; 95% CI: 0·44-0·63; p-value<0.001). Predictors for poor mental health were younger age, food insecurity, water insecurity, lack of legal status documentation, irregular employment, higher intensity of bodily pain, having debt, and having chronic illnesses. The final model demonstrated good discriminative ability and calibration.

**Interpretation:** Mental health predictors were related to basic needs, rights and financial barriers. These allow humanitarian organizations to identify high risk individuals, organizing interventions, and addressing root causes to boost resilience and well-being among older Syrian refugees in Lebanon.

**Funding:** ELRHA’s Research for Health in Humanitarian Crisis Programme.

**Research in context:** 

**Evidence before this study:** A search was conducted on PubMed and Google Scholar for studies published between February 1, 2020 and June 20, 2023, using the search terms “Syrian Refugees”, “Mental Health”, and “Prediction Model”, including all article types with no time constraints or language restrictions. We found that few previous prognostic models for Syrian refugees have been developed exclusively among participants at high risk of poor mental health, such as widowed women, Syrian refugees with post-traumatic stress disorder, or those who experienced ambiguous loss. Older adults were underrepresented in these studies, which had small sample sizes and focused primarily on inter-relational factors. Therefore, their effectiveness in predicting outcomes for this highly vulnerable group, which faces distinct circumstances, may be constrained due to their development based on incomparable samples and contexts. Furthermore, none were developed during the COVID-19 pandemic. Overall, the search highlighted the need for research into the specific vulnerabilities and risk factors for mental health faced by the community of older Syrian refugees in Lebanon, as the existing models do not appear to be applicable to this group.

**Added value of this study:** The study developed a prognostic model to predict the risk of poor mental health amongst older Syrian refugees in Lebanon during the COVID-19 pandemic, using predictors that covered economic, social and health factors. Data were collected using a multi-wave panel study. Most participants had poor mental health that increased over the course of the study. Younger age, food insecurity, water insecurity, lack of legal status documentation, irregular employment, higher intensity of bodily pain, having debt, and having multiple chronic illnesses were predictors of poor mental health. These findings are consistent with previous literature on associations between these vulnerabilities and poor mental health amongst refugees.

**Implications of all the available evidence:** The study provides evidence that the population of older Syrian refugees in Lebanon faces multiple vulnerabilities and were largely at risk for poor mental health, which increased during the COVID-19 pandemic. Vulnerabilities identified in this study as predictors of poor mental health indicate that it will be necessary to engage with humanitarian sectors outside of health such as food assistance, water, sanitation and hygiene (WASH) and legal assistance programs in order to support mental health in older Syrian refugees.

## Introduction

Older Syrian refugees may be at increased risk of experiencing poor mental health due to the exacerbation of stressors during the COVID-19 pandemic.^1^ Those who have experienced war, persecution, displacement, lack of quality shelter, lack of job security and fear of deportation are at an increased risk of mental health problems.^2^ Moreover, the COVID-19 pandemic generated an additional set of challenges for refugees, including difficulties in adhering to preventive measures due to overcrowding, limited access to information and healthcare services, jobs loss and experiencing discrimination and stigmatization, as the host community perceived refugees as a potential source of communicable diseases transmission.^3,4^ These combined stressors may have led to a deterioration in mental health of refugees.^4^

Lebanon hosts approximately 1.5 million Syrian refugees including 831,053 registered with the UNHCR.^5^ A study conducted in Lebanon between 2018 and 2020 showed that the prevalence of moderate to severe depression symptoms among 3,255 Syrian adults was 22%,^6^ which is higher than the depression rate among the host population (9.9%).^7^ Additionally, being 45 years or older was linked to an increased likelihood of experiencing such symptoms.^6^ This figure is likely to have increased due to the co-occurrence of a severe economic crisis, political instability and the pandemic, which has further reduced the limited governmental social protection and mental health services for Syrian refugees, particularly among older individuals and those with mental health disorders.^8^

Older adults were more generally at a higher risk of experiencing the deleterious psychosocial effects of the COVID-19 pandemic.^9^ An analysis of the English Longitudinal Study of Ageing, carried out among 5,146 participants aged at least 50 years old, showed a significant increase in levels of depression and loneliness during the evolvement of COVID-19 pandemic, in contrast to the initial levels reported before the outbreak.^10^ Similarly, a study from the global ApartTogether survey, conducted among 20,742 refugees and migrants, demonstrated that the mental health of older adults was affected during the pandemic.^4^ There are different pathways that could explain this decline in their mental health status, including social isolation, lack of social support, low resilience and the fear of infection.^4,10^

In this context, prediction models have the ability to identify those at high risk of experiencing poor mental health, and informing humanitarian programming for this particularly vulnerable sub-population. To date, previous prognostic models on Syrian refugees have been developed only among very specific subgroups such as widowed women,^11^ those with diagnosed post-traumatic stress disorder,^12^ or those who experienced ambiguous loss,^13^ and are therefore not generalizable to older Syrian refugees. As well, none of these studies were developed during the COVID-19 pandemic.^11–13^ Moreover, there have been calls to concurrently collect data on water security and food security, along with mental health outcomes, to understand the risk each poses to health.^14,15^ In addition, to the best of our knowledge, there are no longitudinal studies assessing the evolution of poor mental health or developing predictive models among older Syrian refugees in the Middle East and North Africa (MENA) region during COVID-19.

Therefore, this study aims to describe the evolution of poor mental health over time and to develop and internally validate a prediction model for poor mental health among older Syrian refugees in Lebanon.

## Methods

### Study design and setting

This study used repeated cross-sectional data from a five-wave study that aimed to examine the vulnerabilities of older Syrian refugees living in Lebanon during the COVID-19 pandemic. This study complied the Transparent Reporting of a Multivariable Prediction Model for Individual Prognosis or Diagnosis (TRIPOD)^16^ and Strengthening the Reporting of Observational Studies in Epidemiology (STROBE)^17^ reporting guidelines for prediction modelling.

### Sampling and study population

Using a list of beneficiaries of a humanitarian non-governmental organization [Norwegian Refugee Council] from 2017-2020, all Syrian refugee households with at least one adult aged 50 years or older were invited to participate (n=17,384). If there was more than one eligible participant in a household, one participant was randomly chosen. Verbal informed consent was obtained from all participants and the data were collected via telephone interviews conducted in Arabic. Participants aged 65 years or older underwent a capacity assessment to consent before participation.^18^ The study population included participants who had complete responses for the mental health outcome measure in the first two waves of the study (September 2020-January 2021), wave three (January-April, 2021) and wave five (January-March, 2022).

### Data sources

The questionnaire for each wave was developed using a combination of sources, including validated questionnaire modules, contextually specific questions, and community-identified priorities. The survey tool was co-created by academics, humanitarian actors, local government officials, and focal points from the refugee communities. It was created in English and translated into Arabic. Modules varied between the waves. The Arabic-language questionnaire was piloted internally with data collectors and local community focal points to ensure face validity. Trained data collectors administered the surveys in Arabic and entered data into structured electronic data collection forms hosted on KoboToolbox. Data entry checks and monitoring were performed for quality assurance throughout data collection.

#### Outcome measure

The primary outcome was poor mental health, which was assessed using the Mental Health Inventory-5 (MHI-5), a subscale of the 36-item Short Form Survey. The MHI-5 is considered as a measure of individuals’ psychological well-being and distress. It includes 5 items assessing the frequency of depressive and anxiety symptoms (being nervous, feeling blue and down), as well as positive signs of mental health (feeling calm and being happy), during the past month. Each item is rated on a 6-point scale, which were summed to produce a total score ranging from 5 to 30, with low values indicative of poor mental health. The MHI-5 had excellent reliability in this population (Cronbach’s alpha=0.82). The total score was transformed using a linear transformation according to standard methods into a score ranging between 0 and 100.^19^ A score of 60 or less, a widely used cut-off point,^19^ indicated poor mental health.

### Candidate predictors

Nineteen possible predictors were identified in the literature and included in the full model including sex, age, education, marital status, living arrangement (living alone vs with others), having unmet waste management needs, number of self-reported physician diagnosed chronic illnesses, intensity of bodily pain measured on a scale from 1 (no pain) to 6 (very severe pain), housing eviction notice, household water insecurity (measured using the short-form Household Water Insecurity Scale),^20^ household food insecurity (measured using the Food Insecurity Experience Scale),^21^ regular employment (if they engaged in paid, regular work in the last 7 days), having received humanitarian cash assistance, having received other kind of assistance, having debt, self-reported experience of any kind of physical abuse or violence inside or outside home, self-reported experience of any kind of verbal abuse or violence inside or outside home, and residence inside or outside informal tented settlements (ITS). Having legal status documentation was also a predictor included in the full model; regularisation of legal status allows refugees to legally remain in Lebanon in case their residency visa expired, or if they entered the country through an unofficial border.

### Missing data

Missing data in item responses were negligible (all variables had <5% missing). Missing responses were assumed to be missing at random and complete case analysis was used.

### Statistical Analyses

Absolute frequencies, proportions and unadjusted logistic regression models with odds ratios (OR) and 95% confidence intervals (95% CI) were presented. Additionally, the growth curve model was used to assess the trend of poor mental health over time across waves one, three, and five. The odds of having poor mental health for each candidate predictor were first examined using bivariate analyses. Data from the first two waves were used for model development.

The variables noted as candidate predictors for mental health outcomes were included in a full multivariable logistic regression model. A stepwise backwards method (p<0·157) was used to remove variables, leaving the model with the best set of predictors for the outcome.^22^ Multicollinearity between variables was tested using a Variance Inflation Factor (VIF) where VIF >5 showed collinearity. The C-Statistic (area under the receiver-operating characteristic curve) was used to assess discrimination in the final model. A C-Statistic value of 1 shows perfect discrimination between individuals who had a poor mental health outcome and those who did not, while a value closer to 0.5 indicates poor discriminative ability.

Calibration plots of the final model were presented. Calibration indicates agreement between the observed and expected outcomes (predictive probabilities) in a model. The plots were created using ten risk groups (grouped based on predictive probabilities). The plots show the mean observed proportion of events against each group’s mean predicted risk of the outcome. Perfect calibration is shown by an intercept of 0 and slope of 1. Overfitting is indicated by a slope <1, which occurs when those with a low risk of the outcome are underestimated, and those with a high risk of the outcome are overestimated. Calibration-In-The-Large (CITL) was used to evaluate the difference between the actual number of individuals with poor mental health and the average predictive risk of this outcome.

The predictors, discrimination, and calibration-slope estimate of the final model were internally validated, using bootstrapping techniques with replacement, where 500 bootstrap samples were used. This generated optimism estimates, an optimism-adjusted C-statistic, optimism-adjusted CITL, and an optimism-adjusted calibration plot. Beta coefficients of the final model were modified using bootstrap shrinkage (multiplying the optimism-adjusted C-slope by the Beta coefficients), and odds ratios were presented. All statistical analyses were undertaken using Stata v.17.

### Ethical approval

Ethical approval for this study was obtained from the American University of Beirut Social and Behavioural Sciences Institutional Review Board [Reference: SBS-2020-0329].

## Results

Out of 3,838 participants who were eligible and consented to participate, 3,322 completed both waves 1 and 2; of those, 3,229 participants had available data on their mental health status (Figure 1). The median age of the study sample was 56 (IQR=53-62; range=50-96); 47.5% were female, 98.1% lived with others, 70.7% had a partner, 3.1% reported physical abuse and 10.9% reported verbal abuse, 92.7% were in debt, 69.8% received cash assistance, 20.1% received other assistance, 29.6% had received an eviction notice, and 47.8% did not report experiencing bodily pain. Most of the participants did not have regularised legal status documentation (75.8%), lived outside ITS (61.2%), had multiple chronic illnesses (43.7%), and 48.4% had not previously attended school. Only 1.9% were regularly employed, 91.9% had experienced household food insecurity, and 30.7% lived in water insecure households (Table 1). In addition, 3,156 and 3,370 participants completed waves 3 and 5 respectively, and had non-missing data on mental health status. The characteristics of these participants are presented in supplementary tables (P2 to P4). Most participants had MHI-5 score ≤ 60 across all waves (76·7% at wave 1, 89·2% at wave 3, and 92·7% at wave 5) and the frequencies demonstrate a statistically significant increase over time (β=0·52; 95% CI: 0·44-0·63; p-value <0.001) (Figure 2).

**Figure 1.**
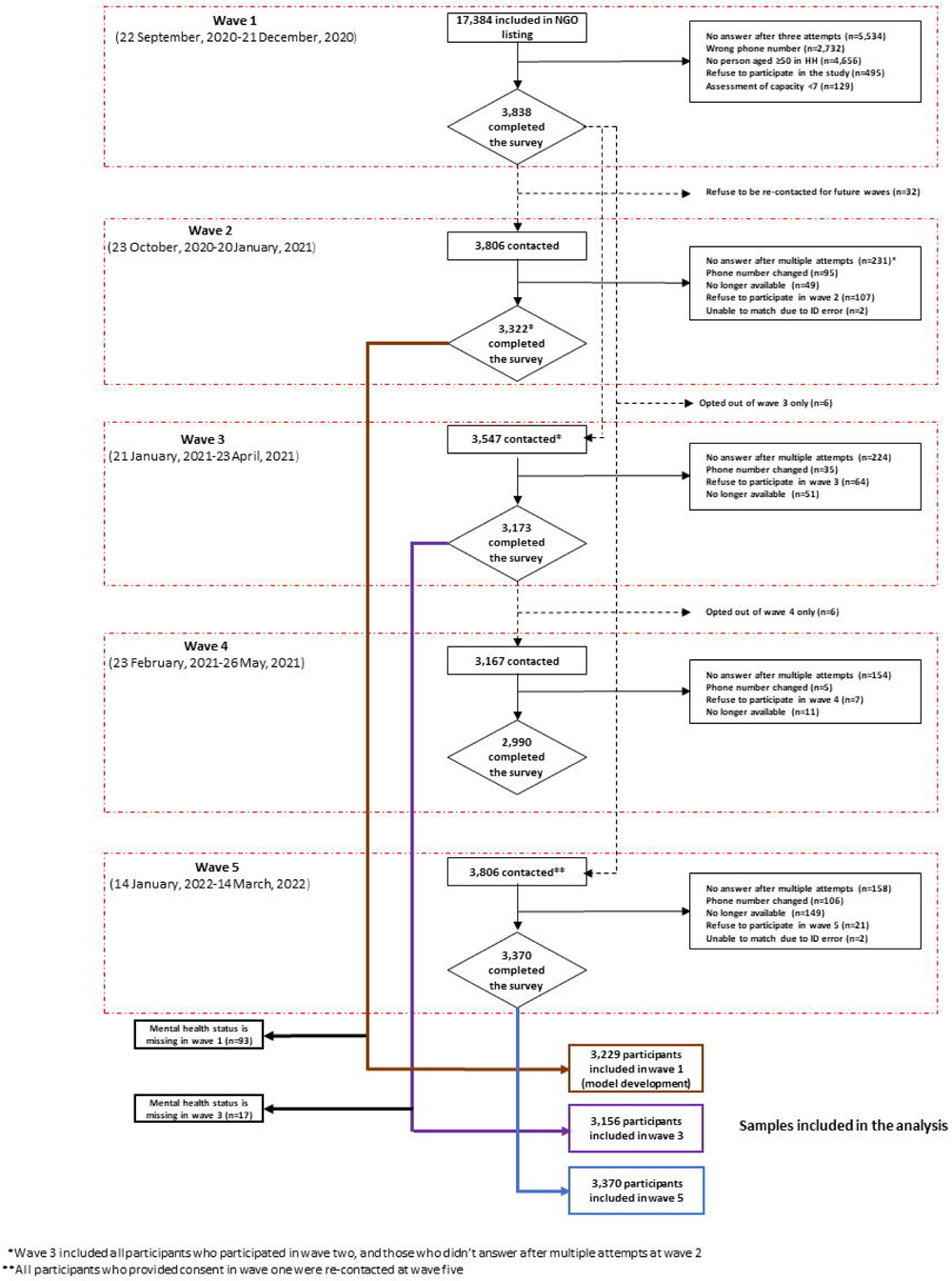
Flow diagram of Syrian refugees included in the study population.

**Figure 2.**
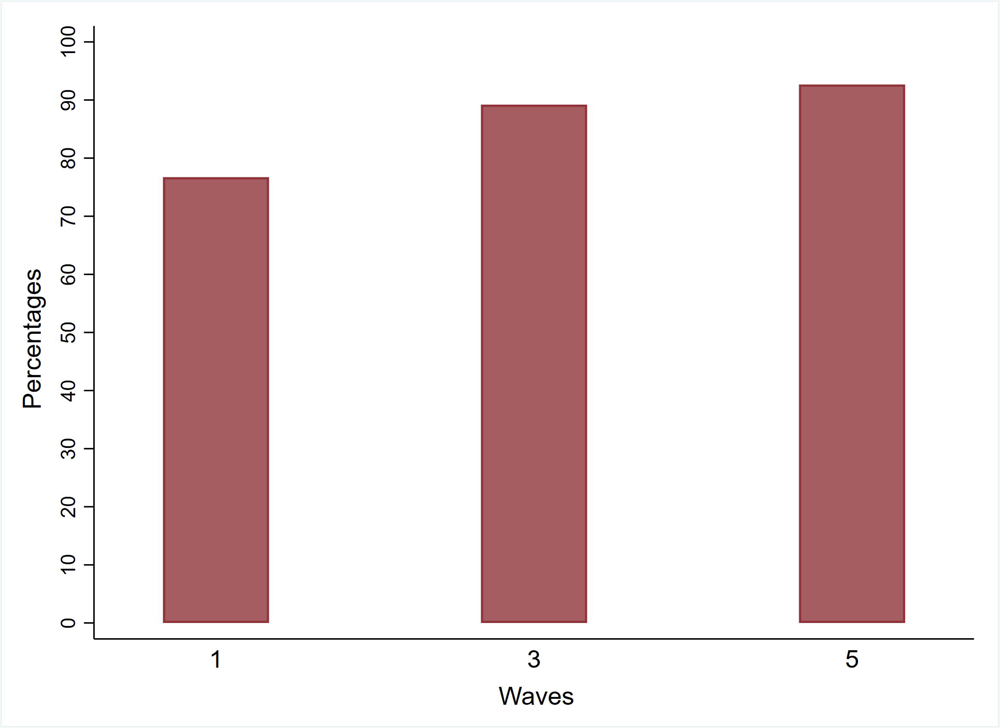
Prevalence of Poor Mental Health across waves 1, 3 and 5.

**Table 1:**
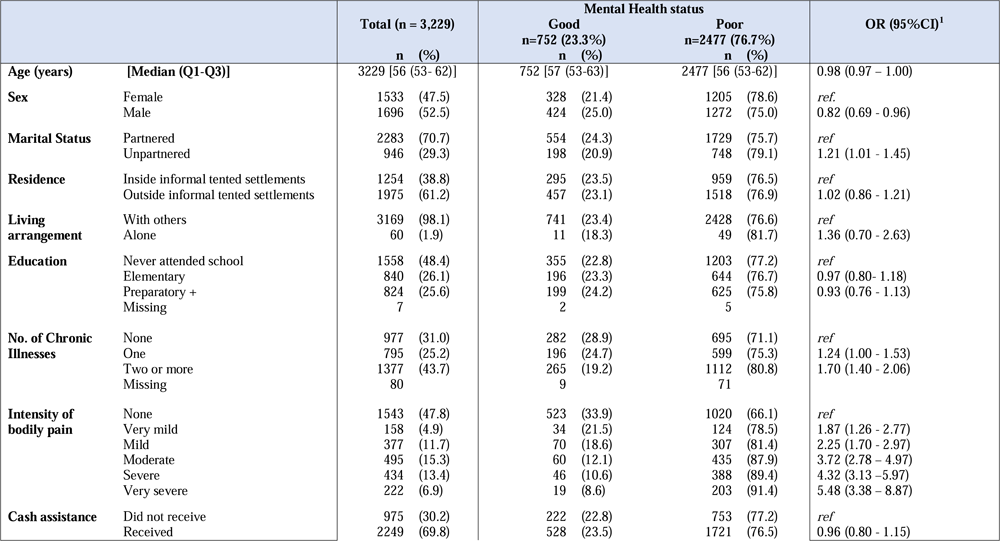
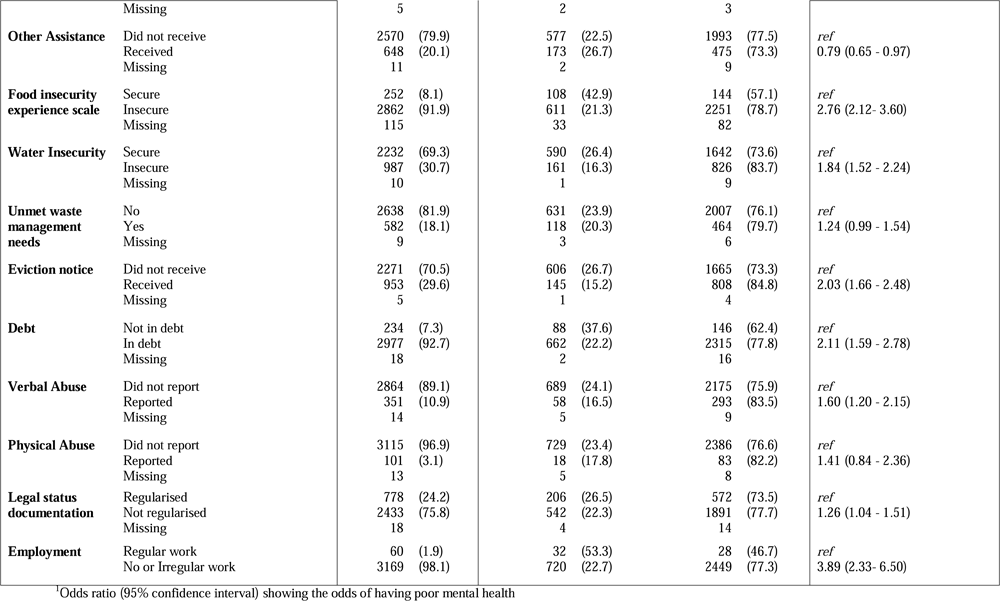
Characteristics of older Syrian refugees and their associations with their mental health status (wave 3)

Unadjusted odds ratios of having an MHI-5 score ≤ 60 are presented for each potential predictor. Characteristics associated with increased odds of having an MHI-5 score ≤ 60 included being unpartnered compared to partnered (OR: 1·21; 95% CI: 1·01-1·45), being food insecure compared to being food secure (OR: 2·76; 95% CI: 2·12-3·60), being water insecure compared to being water secure (OR: 1·84; 95% CI: 1·52-2·24), having two or more chronic illnesses compared to none (OR:1·70; 95% CI: 1·40-2·06), receiving an eviction notice compared to not receiving one (OR: 2·03; 95% CI: 1·66-2·48), being in debt compared to having no debt (OR: 2·11; 95% CI: 1·59-2·78), reporting verbal abuse (OR:1·60; 95%CI: 1·20-2·15) compared to not reporting it, not having regularised legal documentation status (OR:1·26; 95%CI: 1·04-1·51) compared to having regularised status, and having no work or irregular work compared to having regular work (OR: 3·89; 95% CI: 2·33-6·50). Additionally, the participants showed a progressive increase in the likelihood of having an MHI-5 score ≤ 60 with experiencing higher intensity of bodily pain compared to not experiencing it, with OR ranging from 1·87 (95% CI: 1·26-2·77) for very mild pain to 5·48 (95% CI: 3·38-8·87) for very severe pain (Table 1).

The final prediction model for poor mental health (MHI-5 score ≤ 60) retained eight predictors: age, food insecurity, water insecurity, employment status, having debt, legal status documentation, intensity of bodily pain and number of chronic conditions (Table 2). Figure 3 presents the calibration plot adjusted for optimism, whereas the calibration plot of the apparent model before correction for optimism is shown in the appendix (P6). After adjustment for optimism, the final model had a C-statistic of 0·689 (95%CI: 0·667–0·711), which showed good discriminative ability, and a calibration slope of 0·933 (95%CI: 0·821–1·074) as well as CITL of -0·003 (95%CI: -0·082–0·093), indicating a good calibration (Figure 3). The odds ratios along with their coefficients have been adjusted using bootstrap shrinkage and are presented in Table 2.

**Figure 3:**
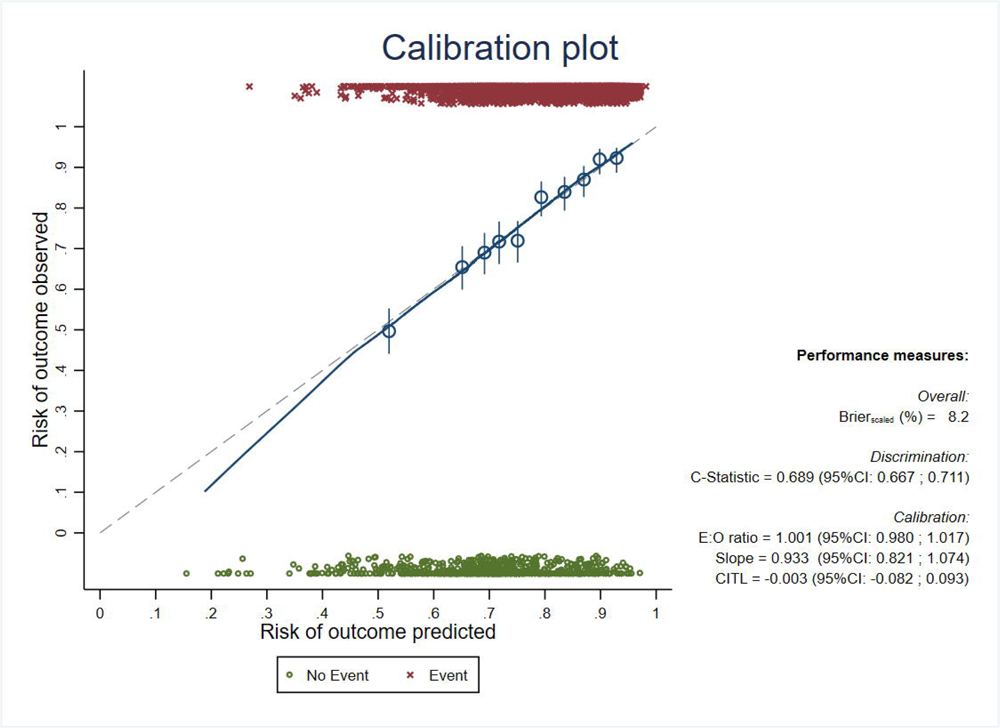
Models performance after adjustment for optimism at wave 1; footnotes: CITL= Calibration in Large; E:O ratio = Expected vs Observed ratio; 95%CI = 95% Confidence interval.

**Table 2:**
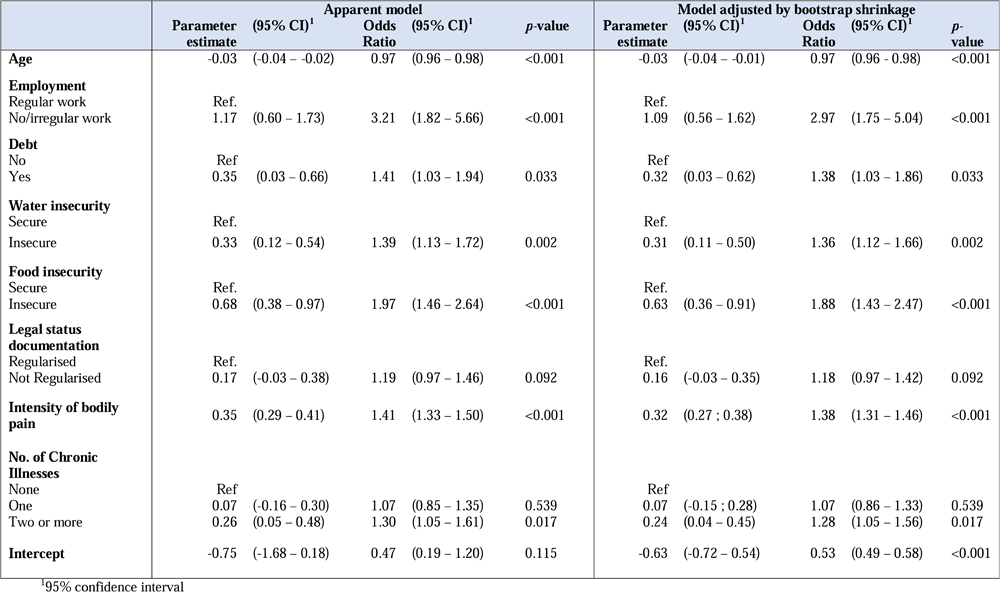
Characteristics of older Syrian refugees and their associations with their mental health status (wave 5)

In the final model, coefficients of predictors indicated the expected directions of association with the outcome. Household food and water insecurity, younger age, higher intensity of bodily pain, having multiple chronic illnesses, having no work or irregular work, having debt and not having regularised legal status documentation increase the likelihood of having a MHI-5 score ≤ 60 (Table 2).

To demonstrate the model, the predicted probability of having an MHI-5 score ≤ 60 (poor mental health), for a 60-year-old Syrian refugee who was not engaging in regular work, had debt, experienced household water insecurity and household food insecurity, lacked regularised legal status documentation, suffered from very severe bodily pain and had multiple chronic illnesses was 90.3%. Meanwhile, a person who was 60 years old, engaging in regular work, not having debt, water secure, food secure, with regularised legal status documentation, not suffering from bodily pain or chronic illnesses had a predicted probability of having a MHI-5 score ≤ 60 (poor mental health) at 10.8%.

## Discussion

This study showed that poor mental health among older Syrian refugees was very common and increased over time during the COVID-19 pandemic in Lebanon. We developed a predictive model of poor mental health that included younger age, food insecurity, water insecurity, lack of legal status documentation, irregular employment, higher intensity of bodily pain, having debt, and having multiple chronic illnesses. This research provides insight into specific socio-ecological vulnerabilities faced by older Syrian refugees (Appendix P7).

Mental health in older Syrian refugees in Lebanon deteriorated over time during the pandemic; this finding is similar to results of the English Longitudinal Study of Ageing and of the global ApartTogether survey.^4,10^ Experiencing daily stressors could have a significant negative impact on mental health^4^ as refugees faced distressing circumstances compounded by social adversity, the economic crisis, and the pandemic.^2,3^

Undocumented older Syrian refugees have higher odds of experiencing poor mental health compared to those with a regularised legal status, which is consistent with other study findings.^23^ Undocumented refugees could face a high burden of anxiety, depression, fear of deportation, lack of social protection and barriers to access healthcare services.^23,24^ In Lebanon, refugee registration was suspended in 2015, at the request of the government. A longitudinal study conducted on 387 migrants in Switzerland demonstrated that regularization had a direct positive impact on reducing the severity of depression.^24^ This indicates that the precarity of legal status needs to be addressed in the context of Syrian refugees in Lebanon.

Food and water insecurity are two distinct but interconnected predictors of poor mental health identified in this study. These types of resource insecurities are well known stressors, linked to worry, distress and increased depressive symptoms, which could affect individuals’ well-being and are both basic needs that intersect.^15^ Water and food insecurities have reciprocal effects, leading to potentially additive and even multiplicative deterioration in mental health status, when experienced together.^25^ It is therefore important for humanitarian interventions aiming to improve mental health in such settings to ensure that refugees receive basic needs support that can alleviate food and water insecurities.

Other predictors for poor mental health that emerged from this study were intensity of bodily pain and having multiple chronic illnesses. The relationships between mental health problems and pain as well as suffering from multiple chronic illnesses such as hypertension and diabetes are well-established in the literature.^26,27^ For instance, complex biological, psychological, and social factors interact and may lead and exacerbate pain and disrupt individuals’ daily life activities,^26^ which increases their need to healthcare services and the use of medication. In particular, older Syrian refugees in Lebanon have reported difficulties in accessing necessary healthcare and medication, factors that are associated with poor mental health among migrants and refugees.^5,28^ Financial factors also exacerbate problems with healthcare access with 60% of Syrian refugees reporting that they reduced healthcare expenditure to be able to cope financially in 2021.^5^

Indebtedness and lack of engagement in regular work are stressors that are related to financial status, which may contribute to poor mental health amongst older Syrian refugees. For most older adults in this study, humanitarian cash assistance was their primary source of income.^5^ Refugees associate emotional well-being with secure employment and the absence of general economic worries.^29^ For instance, Syrian refugees in Lebanon have a lack of employment opportunities at all ages and this is likely to be exacerbated at amongst older adults.

Results of this study demonstrate that older refugees face multiple stressors that contribute to poor mental health. Actually, Syrian refugees referred to environmental and structural stressors, including lack of basic needs, unemployment and movement restrictions as some of the main cause of their emotional distress, which they considered as a normal collective result of pressure accumulation.^30^ These factors are not necessarily recognized by all mental health service providers, though some do recognize that referral to mental health services is sometimes premature considering basic needs are not being sufficiently addressed.^30^

In the context of the pandemic, mental health interventions for older refugees are necessary and need to be linked to the provision of other essential humanitarian services including but not limited to social safety net programs that alleviate food insecurity (e.g., cash interventions, food assistance) water and sanitation interventions and legal and protection services. Predictive models of mental health, could be applied to target individuals at high risk of experiencing mental health problems in order for them to be targeted with interventions aiming to reduce vulnerabilities that impact health and wellbeing.

This study is one of the largest studies to explore the mental health status of older Syrian refugees in Lebanon, with a response rate higher than 85%. Moreover, a limited number of predictors of poor mental health with good predictive abilities were identified in this study; potentially providing a shortlist of predictors to include in data collection efforts of researchers and aid agencies, decreasing intrusiveness and time burden for those surveyed, especially when they are approached by multiple agencies. We acknowledge that the generalizability of these results was limited by the fact that our sample was obtained from a single humanitarian organization’s list of beneficiaries and that several variables rely on self-reported indicators; although this humanitarian organization is one of the largest providers of assistance in Lebanon. Additionally, there are other potential predictors for poor mental health identified in the literature, but not included in our model, such as experiencing trauma during their life course, loss of family members or friends, personal or family history of mental health issues, which could improve the discrimination of our model. In particular, we acknowledge the role of social networks and support as protective factors for mental health. Nevertheless, social support variables were excluded from the model due to their negative impact on the model’s performance.

Future research should evaluate the feasibility and face validity of the proposed predictive tool through qualitative methods, in collaboration with refugees and NGO workers. In addition, we also plan to investigate factors associated with the deterioration of mental health over time to improve the predictive value of these models. In particular, this longitudinal analysis will fill the gap in the literature by examining the causal connections between food and water insecurity, and their intersection with mental health outcomes.^14^

In conclusion, this study indicates that during the COVID-19 pandemic, poor mental health amongst older Syrian refugees in Lebanon may be predicted by age, food insecurity, water insecurity, employment status, having debt, having multiple chronic illnesses, legal status documentation, and intensity of bodily pain. This model can allow humanitarian aid to target those at the highest risk of poor mental health as well as organise interventions that could help older Syrian refugees to meet and maintain their basic human needs and rights, to improve their resilience, health and well-being. In order to holistically address the mental health of this population vulnerabilities that are predictive of poor mental health must be directly addressed, in addition to the provision of mental health services.

## Data sharing statement

The anonymized data can be requested upon reasonable request from NRC and AUB.

## Authors contributions

SA, HG, and SM conceptualized the study and the survey design. SM, HG, SA, BA and TK contributed to data collection. LF contributed to the literature search. BA, LF, and SM drafted the manuscript. The following drafts were reviewed and revised by HG, SM and BA. The underlying survey data were verified by LF, SM and BA. SM supervised LF, BA and TK throughout the project. ZR was involved in the administration and implementation of the study. AMS, CM, ZR and MA contributed to the interpretation of the results. All authors have reviewed and approved the final version of the paper.

## Online Appendix Content

Figure 1: Model performance for the apparent model of wave 1

Figure 2: Timeline of COVID-19 events and the data collection

Table 1: Characteristics of older Syrian refugees and their associations with their mental health status (wave 1)

Table 2: Multivariable model for predicting poor mental health (wave 1)

## Supporting information

Online Appendix Content

## Data Availability

The anonymized data can be requested upon reasonable request from NRC and AUB.

## References

1. Winkler P, Formanek T, Mlada K, et al. Increase in prevalence of current mental disorders in the context of COVID-19: analysis of repeated nationwide cross-sectional surveys. Epidemiology and Psychiatric Sciences 2020; 29: e173.

2. Porter M, Haslam N. Predisplacement and postdisplacement factors associated with mental health of refugees and internally displaced persons: a meta-analysis. Jama 2005; 294(5): 602–12.

3. Bukuluki P, Mwenyango H, Katongole SP, Sidhva D, Palattiyil G. The socio-economic and psychosocial impact of Covid-19 pandemic on urban refugees in Uganda. Soc Sci Humanit Open 2020; 2(1): 100045.

4. Spiritus-Beerden E, Verelst A, Devlieger I, et al. Mental Health of Refugees and Migrants during the COVID-19 Pandemic: The Role of Experienced Discrimination and Daily Stressors. International Journal of Environmental Research and Public Health 2021; 18(12): 6354.

5. UNHCR, UNICEF, WFP. VASyR 2022: Vulnerability Assessment of Syrian Refugees in Lebanon. 2023.

6. Naal H, Nabulsi D, El Arnaout N, et al. Prevalence of depression symptoms and associated sociodemographic and clinical correlates among Syrian refugees in Lebanon. BMC Public Health 2021; 21(1): 217.

7. Karam EG, Mneimneh ZN, Dimassi H, et al. Lifetime prevalence of mental disorders in Lebanon: first onset, treatment, and exposure to war. PLoS Med 2008; 5(4): e61.

8. Gender-Based Violence Information Management System. Ongoing Impact of the compounded crisis (COVID-19, financial and economic crisis) on the GBV [Internet]. 2021 [cited 2022-07-01]. https://reliefweb.int/report/lebanon/lebanon-gender-based-violence-information-management-system-ongoing-impact-compounded.

9. Iob E, Steptoe A, Zaninotto P. Mental health, financial, and social outcomes among older adults with probable COVID-19 infection: A longitudinal cohort study. Proc Natl Acad Sci U S A 2022; 119(27): e2200816119.

10. Zaninotto P, Iob E, Demakakos P, Steptoe A. Immediate and Longer-Term Changes in the Mental Health and Well-being of Older Adults in England During the COVID-19 Pandemic. JAMA Psychiatry 2022; 79(2): 151–9.

11. Hosseini ZA-O, Bakdash T, Ahmad SA-O, Awaad R. Predictors of depression among Syrian refugee women: A socio-culturally relevant analysis. (1741-2854 (Electronic)).

12. Renner A, Jäckle D, Nagl M, et al. Predictors of psychological distress in Syrian refugees with posttraumatic stress in Germany. PLoS One 2021; 16(8): e0254406.

13. Renner A, Jäckle D, Nagl M, et al. Traumatized Syrian Refugees with Ambiguous Loss: Predictors of Mental Distress. Int J Environ Res Public Health 2021; 18(8).

14. Young SL, Bethancourt HJ, Cafiero C, et al. Acknowledging, measuring and acting on the importance of water for food and nutrition. Nature Water 2023; 1(10): 825–8.

15. Young SL, Frongillo EA, Jamaluddine Z, et al. Perspective: The Importance of Water Security for Ensuring Food Security, Good Nutrition, and Well-being. Adv Nutr 2021; 12(4): 1058–73.

16. Collins GS RJ, Altman DG, Moons KG.. Transparent reporting of a multivariable prediction model for individual prognosis or diagnosis (TRIPOD): The TRIPOD statement.

17. Cuschieri S. The STROBE guidelines. Saudi J Anaesth 2019; 13(Suppl 1): S31–s4.

18. Jeste DV, Palmer BW, Appelbaum PS, et al. A New Brief Instrument for Assessing Decisional Capacity for Clinical Research. Archives of General Psychiatry 2007; 64(8): 966–74.

19. Kelly MJ, Dunstan FD, Lloyd K, Fone DL. Evaluating cutpoints for the MHI-5 and MCS using the GHQ-12: a comparison of five different methods. BMC Psychiatry 2008; 8(1): 10.

20. Young SL, Boateng GO, Jamaluddine Z, et al. The Household Water InSecurity Experiences (HWISE) Scale: development and validation of a household water insecurity measure for low-income and middle-income countries. BMJ Global Health 2019; 4(5): e001750.

21. Cafiero C, Viviani S, Nord M. Food security measurement in a global context: The food insecurity experience scale. Measurement 2018; 116: 146–52.

22. Sauerbrei W. The use of resampling methods to simplify regression models in medical statistics. Journal of the Royal Statistical Society: Series C (Applied Statistics) 1999; 48(3): 313–29.

23. Fakhoury J, Burton-Jeangros C, Consoli L, Duvoisin A, Courvoisier D, Jackson Y. Mental health of undocumented migrants and migrants undergoing regularization in Switzerland: a cross-sectional study. BMC Psychiatry 2021; 21(1): 175.

24. Refle JE, Fakhoury J, Burton-Jeangros C, Consoli L, Jackson Y. Impact of legal status regularization on undocumented migrants’ self-reported and mental health in Switzerland. SSM Popul Health 2023; 22: 101398.

25. Young SL, Bethancourt HJ, Frongillo EA, Viviani S, Cafiero C. Concurrence of water and food insecurities, 25 low-and middle-income countries. Bull World Health Organ 2023; 101(2): 90-101.

26. Brooks JM, Polenick CA, Bryson W, et al. Pain intensity, depressive symptoms, and functional limitations among older adults with serious mental illness. Aging & Mental Health 2019; 23(4): 470–4.

27. Sayeed A, Kundu S, Al Banna MH, et al. Mental Health Outcomes of Adults with Comorbidity and Chronic Diseases during the COVID-19 Pandemic: A Matched Case-Control Study. Psychiatr Danub 2020; 32(3-4): 491–8.

28. Farahani H, Joubert N, Anand JC, Toikko T, Tavakol M. A Systematic Review of the Protective and Risk Factors Influencing the Mental Health of Forced Migrants: Implications for Sustainable Intercultural Mental Health Practice. Social Sciences-Basel 2021; 10(9).

29. Noubani A, Diaconu K, Ghandour L, El Koussa M, Loffreda G, Saleh S. A community-based system dynamics approach for understanding factors affecting mental Health and Health seeking behaviors in Beirut and Beqaa regions of Lebanon. Global Health 2020; 16(1): 28.

30. Kerbage H, Marranconi F, Chamoun Y, Brunet A, Richa S, Zaman S. Mental Health Services for Syrian Refugees in Lebanon: Perceptions and Experiences of Professionals and Refugees. Qual Health Res 2020; 30(6): 849–64.

