## Supplementary material for "Predicting poor mental health amongst older Syrian refugees in Lebanon during the COVID-19 pandemic: a multi-wave longitudinal study": Online Appendix Content

### **Table 1: Characteristics of older Syrian refugees and their associations with their mental health status (wave 3)**

| **Characteristic^1^** | | **Total (n = 3,156)** | | **Mental Health status** | | | | | **OR (95%CI)^2^** |
| --- | --- | --- | --- | --- | --- | --- | --- | --- | --- |
|  |  |  |  | **Good**  **n=341(10.8%)** | | | **Poor**  **n=2815 (89.20%)** | |  |
|  |  | **n** | **(%)** | **n** | **(%)** | | **n** | **(%)** |  |
| **Age (years)** | **[Median (Q1-Q3)]** | 3,155 [56 (53- 63)] | | 341 [57 (53-62)] | | | 2814 [56 (53-63)] | | 0.99 (0.98 – 1.01) |
| **Sex** | Female | 1491 | (47.2) | 140 | (9.4) | | 1351 | (90.6) | *ref.* |
|  | Male | 1665 | (52.8) | 201 | (12.1) | | 1464 | (87.9) | 0.75 (0.60 - 0.95) |
| **Marital Status** | Partnered | 2244 | (71.1) | 243 | (10.8) | | 2001 | (89.2) | *ref* |
|  | Unpartnered | 912 | (28.9) | 98 | (10.7) | | 814 | (89.3) | 1.01 (0.79 – 1.29) |
| **Residence** | Inside informal tented settlements | 1190 | (37.7) | 125 | (10.5) | | 1065 | (89.5) | *ref* |
|  | Outside informal tented settlements | 1966 | (62.3) | 216 | (11.0) | | 1750 | (89.0) | 0.95 (0.75 - 1.20) |
| **Intensity of bodily pain** | None | 726 | (23.0) | 124 | (17.1) | 602 | | (82.9) | *ref* |
|  | Very mild | 96 | (3.0) | 15 | (15.6) | 81 | | (84.4) | 1.11 (0.62 – 1.99) |
|  | Mild | 563 | (17.9) | 71 | (12.6) | 492 | | (87.4) | 1.43 (1.04 – 1.95) |
|  | Moderate | 694 | (22.0) | 79 | (11.4) | 615 | | (88.6) | 1.60 (1.18 – 2.17) |
|  | Severe | 843 | (26.7) | 46 | (5.5) | 797 | | (94.5) | 3.57 (2.50 –5.09) |
|  | Very severe | 234 | (7.4) | 6 | (2.6) | 228 | | (97.4) | 7.83 (3.40 – 18.01) |
| **Cash assistance** | Did not receive | 291 | (9.2) | 28 | (9.6) | | 263 | (90.4) | *ref* |
|  | Received | 2861 | (90.8) | 313 | (10.9) | | 2548 | (89.1) | 0.87 (0.58 - 1.30) |
|  | Missing | 4 |  | 0 |  | | 4 |  |  |
| **Food insecurity experience scale** | Secure | 166 | (5.3) | 47 | (28.3) | | 119 | (71.7) | *ref* |
|  | Insecure | 2950 | (94.7) | 290 | (9.8) | | 2660 | (90.2) | 3.62 (2.53- 5.19) |
|  | Missing | 40 |  | 4 |  | | 36 |  |  |
| **Water Insecurity** | Secure | 2365 | (75.1) | 261 | (11.0) | | 2104 | (89.0) | *ref* |
|  | Insecure | 786 | (24.9) | 80 | (10.2) | | 706 | (89.8) | 1.09 (0.84 – 1.43) |
|  | Missing | 5 |  | 0 |  | | 5 |  |  |
| **Other Assistance** | Did not receive | 2816 | (89.3) | 301 | (10.7) | | 2515 | (89.3) | *ref* |
|  | Received | 337 | (10.7) | 40 | (11.9) | | 297 | (88.1) | 0.89 (0.62 – 1.26) |
|  | Missing | 3 |  | 0 |  | | 3 |  |  |
| **Debt** | Not in debt | 106 | (3.4) | 22 | (20.8) | | 84 | (79.2) | *ref* |
|  | In debt | 3040 | (96.6) | 319 | (10.5) | | 2721 | (89.5) | 2.23 (1.38 – 3.62) |
|  | Missing | 10 |  | 0 |  | | 10 |  |  |
| **Verbal Abuse** | Did not report | 2652 | (84.1) | 323 | (12.2) | | 2329 | (87.8) | *ref* |
|  | Reported | 501 | (15.9) | 18 | (3.6) | | 483 | (96.4) | 3.72 (2.29 – 6.04) |
|  | Missing | 3 |  | 0 |  | | 3 |  |  |
| **Physical Abuse** | Did not report | 3019 | (95.8) | 336 | (11.1) | | 2683 | (88.9) | *ref* |
|  | Reported | 134 | (4.2) | 5 | (3.7) | | 129 | (96.3) | 3.23 (1.31 – 7.95) |
|  | Missing | 3 |  | 0 |  | | 3 |  |  |
| **Work** | Yes, regular | 25 | (0.8) | 7 | (28.0) | | 18 | (72.0) | *ref* |
|  | No or Irregular | 3131 | (99.2) | 334 | (10.7) | | 2797 | (89.3) | 3.26 (1.35- 7.85) |
| **Living arrangement** | With others | 3096 | (98.1) | 339 | (10.9) | | 2757 | (89.1) | *ref* |
|  | Alone | 60 | (1.9) | 2 | (3.3) | | 58 | (96.7) | 3.56 (0.87 – 14.66) |

^1^ Information regarding regularisation, unmet waste management needs, number of chronic conditions and eviction notice were not collected in wave 3

^2^Odds ratio (95% confidence interval) showing the odds of having poor mental health

### **Table 2: Characteristics of older Syrian refugees and their associations with their mental health status (wave 5)**

| **Characteristic** | | | **Total (n = 3,370)** | | | **Mental Health status** | | | | | | | **OR (95%CI)^1^** | |
| --- | --- | --- | --- | --- | --- | --- | --- | --- | --- | --- | --- | --- | --- | --- |
|  |  |  |  |  |  | **Good**  **n=247(7.3%)** | | | | **Poor**  **n=3,123 (92.7%)** | | |  |  |
|  |  |  | **n** | | **(%)** | **n** | | **(%)** | | **n** | **(%)** | |  | |
| **Age (years)** | | **[Median (Q1-Q3)]** | 3,370 [58 (54- 64)] | | | 247 [61 (55-66)] | | | | 3,123 [58 (54-64)] | | | 0.98 (0.97 – 0.99) | |
| **Sex** | | Female | 1595 | | (47.3) | 121 | | (7.6) | | 1474 | (92.4) | | *ref.* | |
|  |  | Male | 1775 | | (52.7) | 126 | | (7.1) | | 1649 | (92.9) | | 1.07 (0.83 – 1.39) | |
| **Marital Status** | | Partnered | 2395 | | (71.1) | 158 | | (6.6) | | 2237 | (93.4) | | *ref* | |
|  | | Unpartnered | 975 | | (28.9) | 89 | | (9.1) | | 886 | (90.9) | | 0.70 (0.54 – 0.92) | |
| **Residence** | | Inside informal tented settlements | 1267 | | (37.6) | 103 | | (8.1) | | 1164 | (91.9) | | *ref* | |
|  |  | Outside informal tented settlements | 2103 | | (62.4) | 144 | | (6.8) | | 1959 | (93.2) | | 1.20 (0.92 - 1.57) | |
| **Intensity of bodily pain** | None | | 496 | (14.7) | | 103 | (20.8) | | 393 | | | (79.2) | | *ref* |
|  | Very mild | | 169 | (5.0) | | 12 | (7.1) | | 157 | | | (92.9) | | 3.43 (1.83 – 6.41) |
|  | Mild | | 524 | (15.5) | | 25 | (4.8) | | 499 | | | (95.2) | | 5.23 (3.31 – 8.26) |
|  | Moderate | | 795 | (23.6) | | 32 | (4.0) | | 763 | | | (96.0) | | 6.25 (4.13 – 9.46) |
|  | Severe | | 966 | (28.7) | | 56 | (5.8) | | 910 | | | (94.2) | | 4.26 (3.01 –6.02) |
|  | Very severe | | 420 | (12.5) | | 19 | (4.5) | | 401 | | | (95.5) | | 5.53 (3.32 – 9.20) |
| **Cash assistance** | | Did not receive | 247 | | (7.3) | 21 | | (8.5) | | 226 | (91.5) | | *ref* | |
|  |  | Received | 3114 | | (92.7) | 224 | | (7.2) | | 2890 | (92.8) | | 1.20 (0.75 - 1.91) | |
|  | | Missing | 9 | |  | 2 | |  | | 7 |  | |  | |
| **Food insecurity experience scale** | | Secure | 107 | | (3.2) | 22 | | (20.6) | | 85 | (79.4) | | *ref* | |
|  |  | Insecure | 3224 | | (96.8) | 220 | | (6.8) | | 3004 | (93.2) | | 3.53 (2.17- 5.76) | |
|  |  | Missing | 39 | |  | 5 | |  | | 34 |  | |  | |
| **Water Insecurity** | | Secure | 2076 | | (61.6) | 189 | | (9.1) | | 1887 | (90.9) | | *ref* | |
|  |  | Insecure | 1292 | | (38.4) | 58 | | (4.5) | | 1234 | (95.5) | | 2.13 (1.57 – 2.88) | |
|  |  | Missing | 2 | |  | 0 | |  | | 2 |  | |  | |
| **Unmet waste management needs** | | No | 2464 | | (82.2) | 184 | | (7.5) | | 2280 | (92.5) | | *ref* | |
|  |  | Yes | 533 | | (17.8) | 25 | | (4.7) | | 508 | (95.3) | | 1.64 (1.07 - 1.52) | |
|  | | Missing | 373 | |  | 38 | |  | | 335 |  | |  | |
| **No. of Chronic Illnesses** | | None | 800 | | (26.4) | 72 | | (9.0) | | 728 | (91.0) | | *ref* | |
|  |  | One or more | 911 | | (30.1) | 71 | | (7.8) | | 840 | (92.2) | | 1.17 (0.83 – 1.65) | |
|  |  | Two or more | 1315 | | (43.5) | 68 | | (5.2) | | 1247 | (94.8) | | 1.81 (1.29 - 2.56) | |
|  | | Missing | 344 | |  | 36 | |  | | 308 |  | |  | |
| **Eviction notice** | | Did not receive | 2507 | | (74.6) | 202 | | (8.1) | | 2305 | (91.9) | | *ref* | |
|  | | Received | 855 | | (25.4) | 45 | | (5.3) | | 810 | (94.7) | | 1.58 (1.13 - 2.20) | |
|  | | Missing | 8 | |  | 0 | |  | | 8 |  | |  | |
| **Other Assistance** | | Did not receive | 2904 | | (86.3) | 192 | | (6.6) | | 2712 | (93.4) | | *ref* | |
|  |  | Received | 461 | | (13.7) | 54 | | (11.7) | | 407 | (88.3) | | 0.53 (0.39 – 0.73) | |
|  | | Missing | 5 | |  | 1 | |  | | 4 |  | |  | |
| **Debt** | | Not in debt | 83 | | (2.5) | 10 | | (12.0) | | 73 | (88.0) | | *ref* | |
|  | | In debt | 3261 | | (97.5) | 233 | | (7.1) | | 3028 | (92.9) | | 1.78 (0.91 – 3.49) | |
|  | | Missing | 26 | |  | 4 | |  | | 22 |  | |  | |
| **Verbal Abuse** | | Did not report | 2276 | | (82.0) | 174 | | (7.6) | | 2102 | (92.4) | | *ref* | |
|  | | Reported | 499 | | (18.0) | 17 | | (3.4) | | 482 | (96.6) | | 2.35 (1.41 – 3.90) | |
|  | | Missing | 595 | |  | 56 | |  | | 539 |  | |  | |
| **Physical Abuse** | | Did not report | 2652 | | (95.9) | 186 | | (7.0) | | 2466 | (93.0) | | *ref* | |
|  | | Reported | 112 | | (4.1) | 6 | | (5.4) | | 106 | (94.6) | | 1.33 (0.58– 3.07) | |
|  | | Missing | 606 | |  | 55 | |  | | 551 |  | |  | |
| **Regularisation** | | Regularised | 493 | | (14.7) | 50 | | (10.1) | | 443 | (89.9) | | *ref* | |
|  | | Not regularised | 2864 | | (85.3) | 196 | | (6.8) | | 2668 | (93.2) | | 1.54 (1.11 – 2.13) | |
|  | | Missing | 13 | |  | 1 | |  | | 12 |  | |  | |
| **Work** | | Yes, regular | 24 | | (0.7) | 2 | | (8.3) | | 22 | (91.7) | | *ref* | |
|  | | No or Irregular | 3346 | | (99.3) | 245 | | (7.3) | | 3101 | (92.7) | | 1.15 (0.27- 4.92) | |
| **Living arrangement** | | With others |  | |  | 243 | | (7.4) | | 3063 | (92.6) | | *ref* | |
|  |  | Alone | 3306 | | (98.1) | 4 | | (6.3) | | 60 | (93.8) | | 1.19 (0.43 – 3.30) | |
| **Education** | | Never attended school | 64 | | (1.9) | 124 | | (7.6) | | 1497 | (92.4) | | *ref* | |
|  | | Elementary |  | |  | 65 | | (7.4) | | 809 | (92.6) | | 1.03 (0.75- 1.41) | |
|  | | Preparatory + | 1621 | | (48.2) | 57 | | (6.6) | | 809 | (93.4) | | 1.17 (0.85 – 1.63) | |
|  | | Missing | 9 | |  | 1 | |  | | 8 |  | |  | |

^1^Odds ratio (95% confidence interval) showing the odds of having poor mental health

### **Figure 1: Model performance for the apparent model of wave 1**

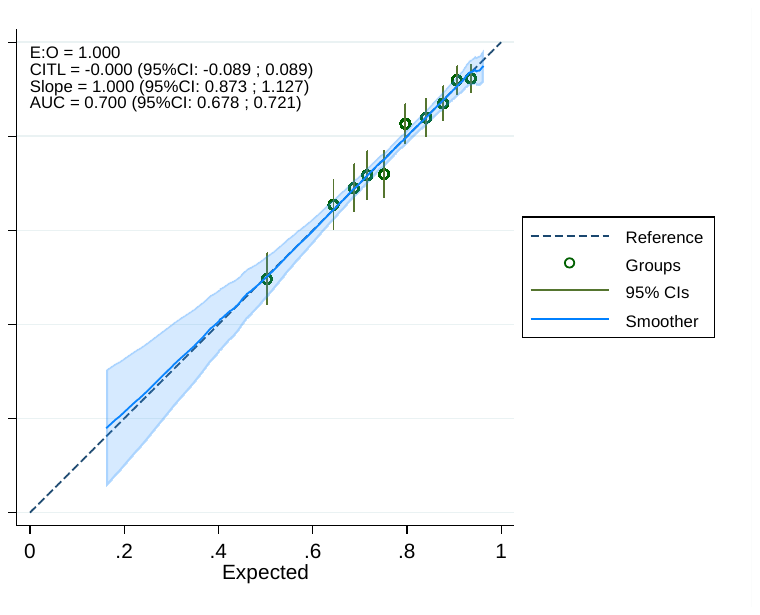

CITL= Calibration in Large

E:0 = Expected to observed ratio

95%CI= 95% confidence interval

AUC: Area Under the Curve

### **Figure 2: Timeline of COVID-19 events and the data collection**

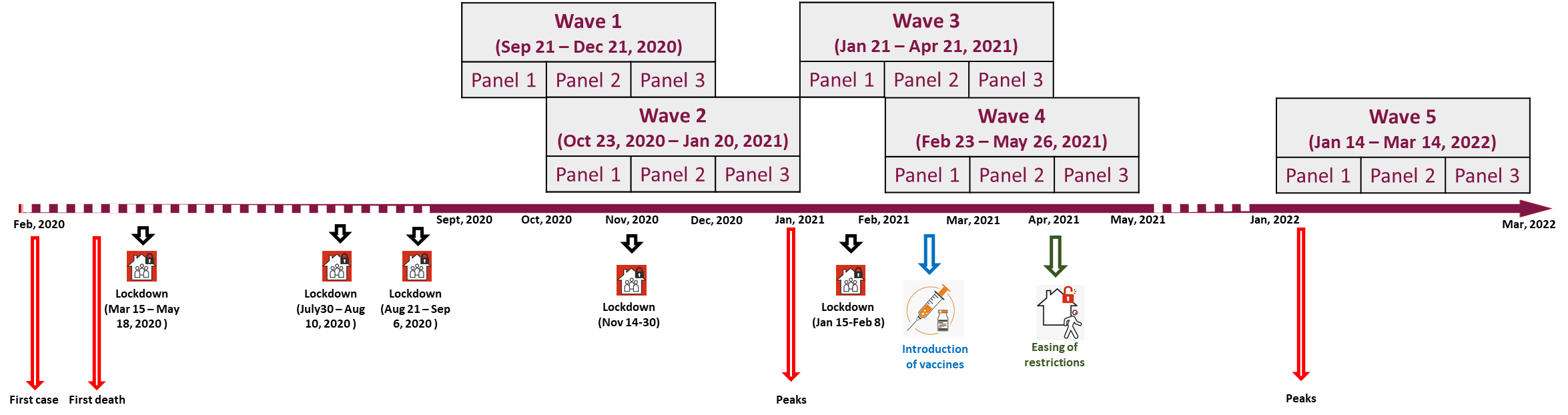
